# A user-centred approach to developing a digital laboratory information system for high-volume clinical data: lessons learned from a malaria study in Kenya

**DOI:** 10.1101/2025.04.24.25326358

**Authors:** Quentin Awori, Brian Polo, Bradley Sandberg, Nickline A. Kuya, Vincent Moshi, Wycliffe Odongo, Daniel Musungu, Kephas Otieno, Mary Omwalo, Nicole L. Achee, John P. Grieco, John E. Gimnig, Eric Ochomo

## Abstract

**Introduction:** Digital health interventions can potentially improve research-related procedures and outcomes. However, many fall short of the expected benefits. Implementing user-centred design (UCD) in their development increases their relevance and adoption. Here we describe a UCD approach in building a data collection platform for use in a research trial requiring timely management of malaria microscopy results.

**Methodology:** UCD was used to transform a paper-based laboratory information system used to manage malaria microscopy to an end-to-end digital equivalent. The approach was based on the three cycles of the information systems research framework i.e. relevance, prototype design and rigour. In the relevance cycle, the paper-based workflow was evaluated and a needs assessment conducted. In the prototype design cycle, a set of electronic case report forms, a revised laboratory request form and an online data dashboard were developed. In the rigour cycle, interviews were held with the relevant users to gain insight on design of each prototype iteration. All staff were trained and the deployment of the finalised digital platform was launched at a single point in time. Three months after implementation of the finalised system, users’ experiences were assessed.

**Results:** Turnaround time on reporting microscopy results reduced from a median of 15 days to 3 days(p value < 0.0001). The amount of paper used was reduced to eight times less than before. Redundant data entry activities were removed, and labour-intensive clerical activities were replaced with automated digital processes. Users reported improved work productivity and efficiency in managing microscopy results.

**Conclusion:** The UCD data platform enabled the design of a contextually relevant, successfully adopted and clinically impactful laboratory information system. The manner in which digital health data collections systems are developed, and introduced to users is as critical as their features and functions

**What is already known on this topic:** Successful deployments of digital health interventions are hard won. Better approaches are needed to develop and deploy them.

**What this study adds:** User centred design approaches can be successfully applied even in the context where the implementation of clinical trials may benefit from a digital health intervention.

**How this study might affect research, practice or policy:** We demonstrate how user centred design was applied. This could serve as a reference for applying it in similar contexts.

## BACKGROUND

Digital health interventions have the potential to improve research-related procedures and outcomes. Such interventions have been used in managing research data generated during large-scale clinical trials with success ^1^. However, many fall short of all the benefits expected from them and are often not adopted or abandoned early in their implementation^2^. Better approaches to developing and deploying digital health applications would increase their relevance and adoption, justifying continued investment. Adopting user-centred design (UCD) approaches is key in the development of such implementations. UCD is a design framework that iteratively studies, designs and evaluates systems by involving the end users and other stakeholders throughout the life cycle^3^. This enhances user acceptance and success when implemented. Here we describe the UCD approach applied to transition from a paper-based laboratory information system (LIS) to an open-source based digital platform for a large-scale clinical trial.

## METHODOLOGY

### Context

The implementation was conducted within the context of a large-scale, cluster randomised, controlled trial evaluating the efficacy of a spatial repellent mosquito vector control intervention to reduce the incidence of malaria in Busia county, Kenya ^4^. Each day, 100-140 blood samples were collected from study participants for use in making blood smears on slides to check for malaria infection by microscopy. Each microscopy slide had two readings specified as Read 1 (R1) and Read 2 (R2), conducted by two independent microscopists. The presence/absence of *Plasmodium* spp. parasites (which is the cause of malaria disease), the type of species, and the parasite density were determined during each R1 and R2. In the event of discordance, an independent microscopist conducted a third reading, Read 3 (R3) on the same blood slide with the outcomes determined by the agreement with either R1 or R2. Study subjects found to have a positive malaria infection were treated with artemether /lumefantrine(AL) at the appropriate dose for parasite clearance according to standard clinical management procedures. Reducing the time between blood sample collection and when test results were ready was important for clinical management of the subject. For the first four months of the study, laboratory data collection consisted of a paper-based data collection system to document blood sample collection. Due to the major delays in obtaining microscopy results, transition to a digital laboratory information system (LIS) was considered.

### Digital Data Collection Platform And Dashboard

Previous to the development of the digital LIS, most of the data collection for other aspects of the study was already using the CommCare© platform via electronic case reports forms (eCRFs). Additionally, a web -based dashboard used to monitor study implementation was in place. It consisted of a cloud-server backend, developed in the Python programming language, using the Django framework, and a PostgreSQL database management system for data storage. The frontend of the dashboard was developed in JavaScript, using the VueJS framework.

### User-Centred Design (UCD) Approach

Our UCD approach was based on the three cycles of information system research framework ^5^: 1) relevance, 2) prototype design and 3) rigour cycles:

<u>1. Relevance cycle:</u> To become more relevant and contextual to the environment in which we were designing the platform we: i) evaluated the current paper-based workflow of the staff cadres that used it most frequently i.e. the clinical team and the laboratory microscopy team; and ii) conducted a needs assessment to understand why paper had historically been used and the concerns different stakeholders wanted addressed in the transition to a digital platform. The stakeholders included senior level malaria laboratory management, the clinical team, laboratory microscopists, research regulatory associates and the quality assurance teams.
<u>2. Prototype design cycle:</u> In this cycle, three components were developed. First, a set of electronic case report forms (eCRFs) for data collection to document sample collection at the clinic, reception of slides transported to the laboratory and microscopy results. This development was guided by the paper forms that were previously used. Because the CommCare© platform, an open-source electronic data collection platform, was already being used in the study for other purposes, it was chosen as the platform for data collection. Second, a new paper-based lab request form (LRF) was developed, to keep track of the batches of slides as they were moved from the health facilities to the laboratory. Third, an online dashboard was developed for the laboratory manager (LM). Using this dashboard, the LM would query various details on any malaria slide including its collection date, the participant it was sampled from and other result details (R1, R2, parasite species).
<u>3. Rigour cycle:</u> With each design iteration of the three prototypes, interviews were held with the relevant users. Responses and comments were used to improve the prototypes. Several iterations of this kind took place until no further design improvements needed to be incorporated.

### User acceptance survey

After approximately five months of using the digital LIS, users (clinical officers and laboratory microscopists) were invited to complete a self administered questionnaire online to understand the modified experiences. The questions were based on the unified theory on the acceptance and user of technology (UTUAT)^6^.

## RESULTS

### Needs assessment of the current paper-based workflow

Using paper forms for data collection had a history of reliability in documenting and record keeping for other large studies conducted by KEMRI. Such paper-based data management systems predated any implementation of a digital LIS platform, and hence changing to the latter had previously been met with hesitancy by staff. Paper forms provided assurance of tangible retrieval for audit needs. The use of organised paper-based data collection systems had matured in the organisation over the previous decades, making them easy to deploy on any new project.

Evaluation of the paper-based data management system determined that the labour intensive clerical activities were the rate limiting factor which led to delays in reporting microscopy results to clinicians. Manual verification by laboratory staff of correct documentation of the subjects’ results, which included verification of participant details and mathematical calculations, had a high cognitive load as it involved attention to detail and calculations of the parasitemia based on the number of parasites counted during microscopy.

Data entry was also very time consuming. To enter the results, each record needed to be searched on the CommCare© platform, which often had > 10,000 cases pending data entry. This search process alone would often take > 60 seconds, and at times fail, needing the tablet to be rebooted. It is our presumption that the O(n) time and memory complexity of this search step did not scale with the high number of cases on the CommCare© platform. This slow processing of paper LRFs resulted in them collecting into stacks, several feet high, occupying considerable workspace in the laboratory.

### Prototype design and improvement

The underlying principle in the design of the prototype platform was to reduce the reliance on paper forms, remove redundant activities, and replace clerical labour-intensive activities with automated digital processes, all the while addressing stakeholder needs (Table 1).

**Table 1:**
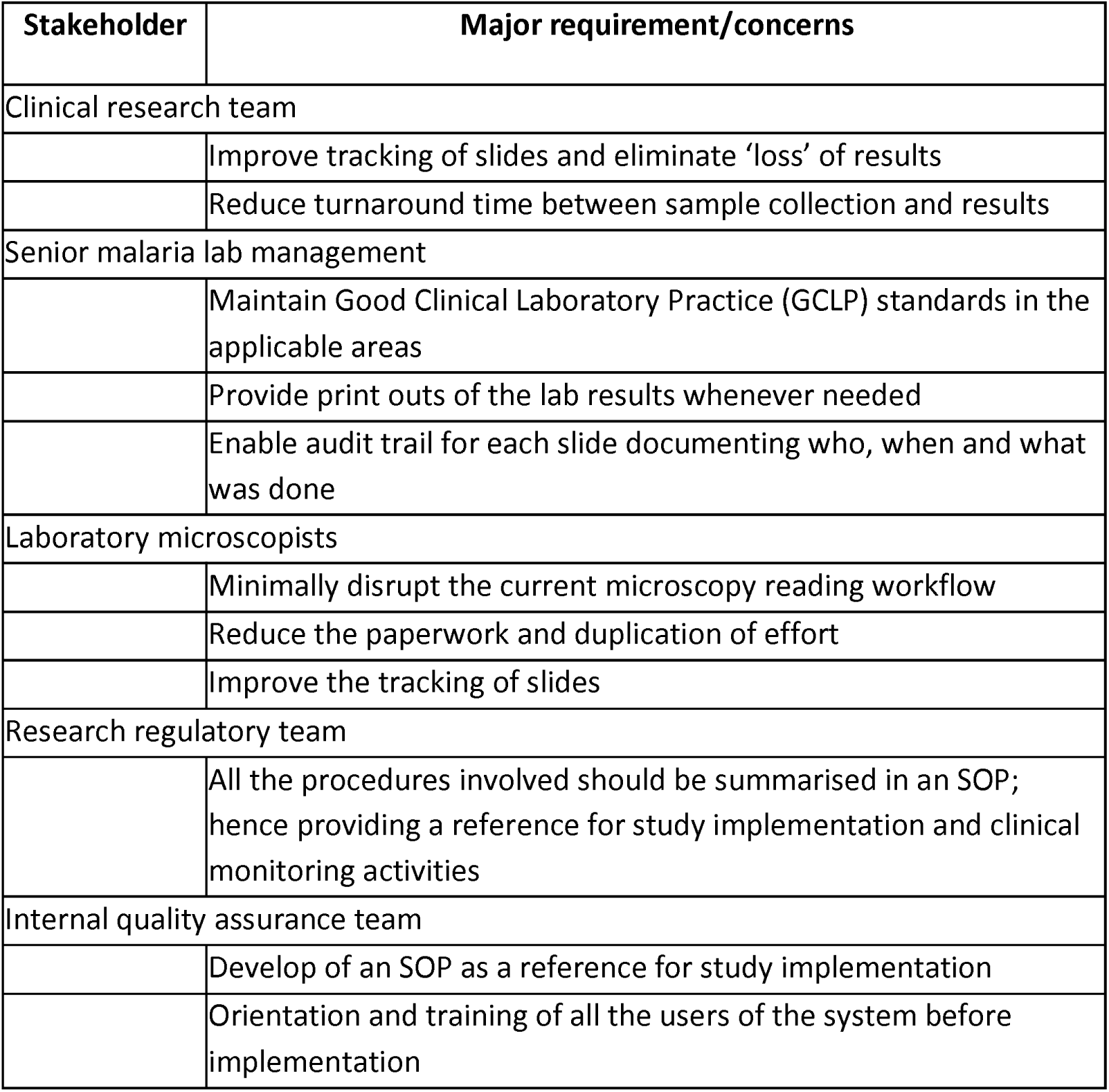
Summary of needs assessment outcomes according to stakeholder category.

Upon evaluation, many parameters that were collected were found to either be unnecessary to the study or redundant having been recorded elsewhere. An abbreviated version of the LRF (SI1) was developed that was faster to complete and reduced the pieces of paper used by a factor of eight compared to the old version. The design of the new LRF was consultative and user centred. Iterative designs involved the input of the LM, clinical officers, data managers, and the clinical quality assurance teams. Importantly, the abbreviated paper form allowed for scannable barcodes to be stuck on (the second column) thereby linking subject metadata with malaria slide results. The barcode was also scanned into the CommCare© platform for referencing. This reduced the likelihood of human data entry error and enabled data set completion checks. These barcodes were attached on the malaria slides and the new LRFs.

At the clinics, information on each malaria slide was entered onto tablets using a sample collection electronic case report forms (eCRFs). This sample collection eCRF incorporated data validation rules, and skip-logic patterns to enhance data quality. The barcodes attached in the malaria slides were also scanned in the eCRF, providing an at-source reference with paper LRFs.

Intentionally, the laboratory’s microscopy reading workflow was not substantively changed. For example, counting of parasites involved the use of a hand-held tally counter as before. The most important change was that the microscopists would now enter the result directly onto a tablet as opposed to the paper forms. To mitigate data entry errors, parameters from the microscopy reading were entered in two iterations of the same set of questions. Validation was then performed automatically by ensuring the responses matched. Additionally, mental calculations that were initially performed manually were now automated, eliminating the cognitive load experienced in the paper system.

The web-based component of the LIS (SI2) was developed for the LM and administrative staff to filter and view status of microscopy data for enhanced data monitoring. Noting that the dashboard would most frequently be used via a tablet, various design features were incorporated to enhance the user experience while using the portal e.g:

i. reducing the click distance for various tasks before a desired result;
ii. using font colours in the rows of data to hint at the status of a slide; black for pending review, green for approved and red disapproved. This was preferred to additional columns showing these statuses, reducing the width of the table and amount of text presented to the user;
iii. minimising typing by categorising the most frequent comments into list box options;
iv. considering finger width and clickable pixel area on the controls - making them large enough even on the tablet interface;
v. enabled searching on various sample metadata such as date of sample collection, status of microscopy R1, R2, R3, and microscopist assignments.

### Training

To facilitate proper use of the new platform, all relevant staff were trained. An SOP offering guidelines on the implementation of the digital LIS was developed and shared with staff members for review and consensus. Clinical officers, lab microscopists and managers were then trained on this SOP before it was deployed. Various recommendations resulting from the training were incorporated in iterative updates before finalisation of the SOP. This training offered an opportunity to address reasons for hesitancy in changing over to the new system.

### Deployment

The deployment strategy depended on whether more staff could be hired at the central lab during the changeover period. As this was not possible, only one system could be implemented at a time. Hence, the recreate (all at once) deployment strategy was chosen; all health facilities and the central lab switched to the digital LIS at the same time. However, users were allowed to revert back to the paper-based forms, if deployment of the digital LIS failed.

The deployment of the digital LIS was uneventful; no hitches or mishaps were experienced. Of note, there was a drastic reduction in necessary work effort experienced by all clinical and laboratory staff.

### Reporting time

As this was important to the study implementation, we evaluated the difference in turnaround time between the paper based and the digital LIS. For each sample, we defined this as the number of days between sample collection and its first reading (R1). The turnaround times in both phases were not normally distributed (Figure 2). The median turnaround time in the paper based system was 15 days (IQR 6-26 ) while that in digital LIS was 3 days (IQR 2-5). This difference was statistically significant (Mann-Whitney U: p value < 0.0001).

**Figure 2:**
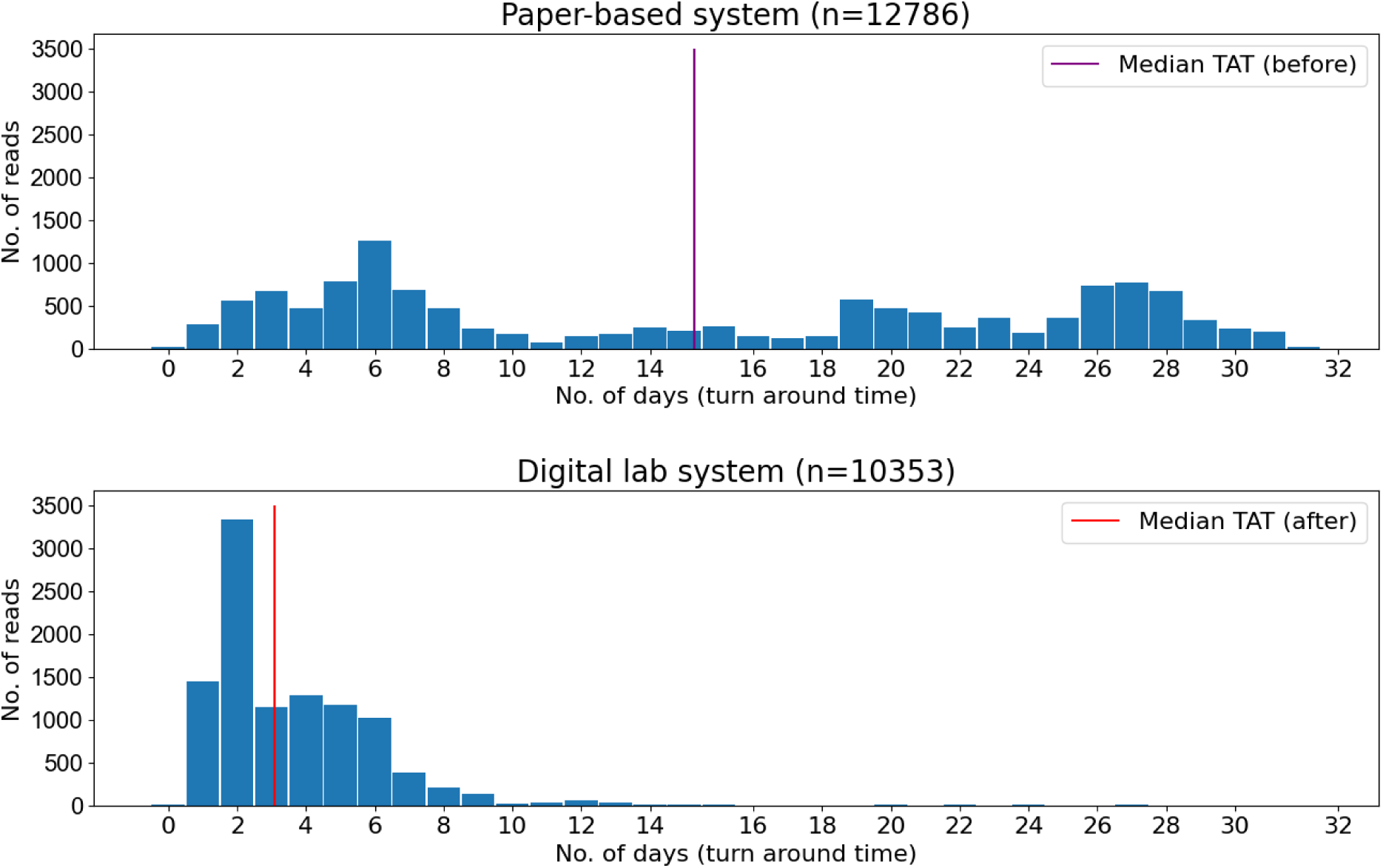
Turnaround time in the paper based (top) and the digital based systems.

### Feedback from Study Staff (Users)

Twenty of 24 staff (83.3%) provided feedback on the UCD platform. 16/20 (80%) at least agreed that the new system improved their work productivity; 16/20 (80%) also agreed that they were more efficient in managing/tracking microscopy results. 17/20 (85%) responded that the new system was easier to learn and use. While 14/20 (70%) stated they would recommend the digital LIS for use in a similar study, the remaining six (30%) indicated they would recommend the digital LIS, but with some modification. As an optional question, we asked staff what further improvements are needed, to which three responses (all from microscopists) suggested the ability to edit microscopy results after they have been entered, in the event of data entry error - while data entry errors on the new system were rare, they had to be corrected via email correspondence with the data team.

## DISCUSSION

Our UCD approach facilitated the development of a contextually relevant, clinically impactful platform in a high data volume clinical research setting. The turnaround time for malaria microscopy results was significantly reduced from a median of 15 to 3 days. User acceptance and adoption were also high.

Our approach was based on the three cycles of the Information System Research framework ^5^. During the relevance cycle, we engaged with staff to understand their concerns, and requirements which bestowed confidence - hesitancy toward digital transformation in healthcare is a known phenomenon ^7,8,9,10^. While this has not been fully understood, it is not entirely unfounded. Even in developed setups, successful implementations of digital health are hard won ^9^. Indeed, the manner in which digital health interventions are implemented is as important as the features and functions of the interventions themselves ^10^.Additionally, during this relevance cycle, we also learned of previous attempts to digitise the LIS at KEMRI which had not been successful. During those past efforts, a UCD approach was not followed.

Staff acceptance and adoption were high. Clinical officers and microscopists were most closely involved in our UCD approach given their roles in data capture. For this reason, it was important these staff members were empowered to perform their duties in an enhanced manner. Indeed, the unified theory for the acceptance and use of technology (UTAUT)^6^ underlines performance expectancy and effort expectancy as important considerations in evaluating the acceptance and use of technology.

We used the recreate deployment strategy, an all at once approach, to changeover to the new system. Other transition strategies would have required double documentation data capture on both paper and the digital LIS, with a potential negative impact on adoption. Double documentation has been indicated as a challenge to adoption of technologies in health systems^11,12^.

There was a significant reduction in data turnaround time, following adoption of the digital LIS. With the previous paper-based system, data within the LRF denoted when a microscopy reading was performed, not when clinical officers obtained the results. In practice, availability of microscopy results had been delayed by several days or weeks after slide reading was completed due to a backlog in data entry and the clerical task of organising the LRFs for each respective clinic. With the digital LIS, microscopy results were immediately available to the clinics, via the internet and the clinicians had results within minutes of the microscopist reading the slide. This improved clinical management of study participants, especially those who were asymptomatic during follow-up visits, as they could be followed up at home promptly in case microscopy results indicated a positive infection for malaria.

While the UCD approach described here was adopted from a well-documented approach ^5^ the generalizability of the LIS platform may be limited when deployed in other settings as it was contextualised for the Kenya study location and specific trial design. The number of staff using the LIS was relatively small (n=24), all of whom were required to use the system in a mandatory rather than voluntary context. Much as staff were assured that their feedback during the UCD approach was anonymised, these factors could have biased their responses towards the acceptance of the digital platform.

## CONCLUSION

UCD approaches should guide the implementation of digital health interventions, or their improvement where they already exist. This would help realise the benefits hoped from them, increase their adoption, utility and sustained use. Keen attention should be paid to the most frequent users and/or use cases anticipated during implementation.

## Supporting information

SI Annex 1

SI Annex 2

## Data Availability

All data produced in the present study are available upon reasonable request to the authors

## ACKNOWLEDGEMENTS

We gratefully thank the study participants and members of study communities in Busia county, Kenya. Special gratitude to study staff and supervisors who worked tirelessly to supervise, implement and/or manage trial activities including data management. We sincerely appreciate staff patience and good cheer adapting to the digital LIS platform as well as Bradley Sandberg, Matthew Noffsinger and the team at the University of Notre Dame Center for Research Computing for developing the custom-designed CommCare© database, assurances of functionality and technical support.

## AUTHOR INFORMATION

### Authors and affiliations

Kenya Medical Research Institute - Centre for Global Health Research, Kisumu *Quentin Awori, Brian Polo, Nickline A. Kuya, Vincent Moshi, Daniel Musungu, Kephas Otieno, Mary Omwalo, Eric Ochomo*

Center for Research Computing, University of Notre Dame, Notre Dame, IN, USA *Bradley Sandberg*

Department of Biological Sciences, Eck Institute for Global Health, University of Notre Dame, Notre Dame, IN, USA

*Nicole L. Achee, John P. Grieco*

Division of Parasitic Diseases and Malaria, Center for Disease Control and Prevention, Atlanta, GA, USA

*Wycliff Odongo, John E. Gimnig*

### Author contributions

All authors contributed to reviewing the manuscript.

QA: conceptualization, data collection, data curation, methodology, data analysis, writing– original draft, developed questionnaire; BP: developed questionnaire, data collection, data analysis, writing–review, project administration; BS: conceptualization, project administration, writing–review; NAK: miscellaneous activities, writing–review, project administration;

VM: data collection, data analysis, writing–review; WO: data collection, writing–review; DM: supervision, project administration, data collection; KO: project administration, supervision;

MO: supervision, miscellaneous activities; NLA: writing–review & editing, supervision, project administration; JPG: writing–review & editing, supervision, project administration; JEG: writing–review & editing, supervision, project administration; EO: conceptualization, writing–review, supervision, project administration.

### Corresponding author

Quentin Awori

## DISCLAIMER

The findings and conclusions in this manuscript are those of the authors and do not necessarily represent the official position of the Kenya Medical Research Institute, the US Centers for Disease Control and Prevention or Unitaid.

## LIST OF ABBREVIATIONS

LIS: Laboratory information system
SOP: standard operating procedure
UCD: user-centred design
UTAUT: Unified theory for the acceptance and use of technology

## REFERENCES

1. Elson WH, Kawiecki AB, Donnelly MAP, et al. Use of mobile data collection systems within large-scale epidemiological field trials: findings and lessons-learned from a vector control trial in Iquitos, Peru. BMC Public Health. 2022;22(1):1924. doi:10.1186/s12889-022-14301-7

2. Greenhalgh T, Abimbola S. The NASSS Framework - A Synthesis of Multiple Theories of Technology Implementation. Stud Health Technol Inform. 2019;263:193–204. doi:10.3233/SHTI190123

3. Cornet VP, Toscos T, Bolchini D, et al. Untold Stories in User-Centered Design of Mobile Health: Practical Challenges and Strategies Learned From the Design and Evaluation of an App for Older Adults With Heart Failure. JMIR MHealth UHealth. 2020;8(7):e17703. doi:10.2196/17703

4. Ochomo EO, Gimnig JE, Bhattarai A, et al. Evaluation of the protective efficacy of a spatial repellent to reduce malaria incidence in children in western Kenya compared to placebo: study protocol for a cluster-randomized double-blinded control trial (the AEGIS program). Trials. 2022;23(1):260. doi:10.1186/s13063-022-06196-x

5. Hevner AR. A Three Cycle View of Design Science Research. 2007;19.

6. Venkatesh V, Morris MG, Davis GB, Davis FD. User Acceptance of Information Technology: Toward a Unified View. MIS Q. 2003;27(3):425–478. doi:10.2307/30036540

7. Cho Y, Kim M, Choi M. Factors associated with nurses’ user resistance to change of electronic health record systems. BMC Med Inform Decis Mak. 2021;21(1):218. doi:10.1186/s12911-021-01581-z

8. Safi S, Thiessen T, Schmailzl KJ. Acceptance and Resistance of New Digital Technologies in Medicine: Qualitative Study. JMIR Res Protoc. 2018;7(12):e11072. doi:10.2196/11072

9. Asthana S, Jones R, Sheaff R. Why does the NHS struggle to adopt eHealth innovations? A review of macro, meso and micro factors. BMC Health Serv Res. 2019;19:984. doi:10.1186/s12913-019-4790-x

10. Ross J, Stevenson F, Dack C, et al. Developing an implementation strategy for a digital health intervention: an example in routine healthcare. BMC Health Serv Res. 2018;18(1):794. doi:10.1186/s12913-018-3615-7

11. Dijkstra A, Heida A, Rheenen PF van. Exploring the Challenges of Implementing a Web-Based Telemonitoring Strategy for Teenagers With Inflammatory Bowel Disease: Empirical Case Study. J Med Internet Res. 2019;21(3):e11761. doi:10.2196/11761

12. Dinh N, Agarwal S, Avery L, et al. Implementation Outcomes Assessment of a Digital Clinical Support Tool for Intrapartum Care in Rural Kenya: Observational Analysis. JMIR Form Res. 2022;6(6):e34741. doi:10.2196/34741

