## Supplementary material for "A user-centred approach to developing a digital laboratory information system for high-volume clinical data: lessons learned from a malaria study in Kenya": SI Annex 1

**Facility:** ______________________________

**Collection Date:**_____/______/________

### SPATIAL REPELLENT STUDY
#### Malaria Microscopy Slide Transfer Log

| PARTICIPANT ID | BARCODE ID | AGE<br>SEX | VISIT TYPE | Expedited<br>Time Collected | Test<br>Staff Initials | Slides Received ☒ |
|---|---|---|---|---|---|---|
| | (Place label here) | Age:<br>☐ M<br>☐ F | | Expedited<br>Yes ☐ No ☐<br>____: ____hrs | Microscopy and Archival ☐ | Slide 1 ☐<br>Slide 2 ☐ |
| | (Place label here) | Age:<br>☐ M<br>☐ F | | Expedited<br>Yes ☐ No ☐<br>____: ____hrs | Microscopy and Archival ☐ | Slide 1 ☐<br>Slide 2 ☐ |
| | (Place label here) | Age:<br>☐ M<br>☐ F | | Expedited<br>Yes ☐ No ☐<br>____: ____hrs | Microscopy and Archival ☐ | Slide 1 ☐<br>Slide 2 ☐ |
| | (Place label here) | Age:<br>☐ M<br>☐ F | | Expedited<br>Yes ☐ No ☐<br>____: ____hrs | Microscopy and Archival ☐ | Slide 1 ☐<br>Slide 2 ☐ |
| | (Place label here) | Age:<br>☐ M<br>☐ F | | Expedited<br>Yes ☐ No ☐<br>____: ____hrs | Microscopy and Archival ☐ | Slide 1 ☐<br>Slide 2 ☐ |
| | (Place label here) | Age:<br>☐ M<br>☐ F | | Expedited<br>Yes ☐ No ☐<br>____: ____hrs | Microscopy and Archival ☐ | Slide 1 ☐<br>Slide 2 ☐ |
| | (Place label here) | Age:<br>☐ M<br>☐ F | | Expedited<br>Yes ☐ No ☐<br>____: ____hrs | Microscopy and Archival ☐ | Slide 1 ☐<br>Slide 2 ☐ |
| | (Place label here) | Age:<br>☐ M<br>☐ F | | Expedited<br>Yes ☐ No ☐<br>____: ____hrs | Microscopy and Archival ☐ | Slide 1 ☐<br>Slide 2 ☐ |

TECH/STAFF SENDING THE SPECIMEN… ............................................Date/Time released: Date______/_______/______Time___ :____hrs

COURIER/DRIVER.......................................................................Date /Time of arrival: Date_______/_____/_________Time____: ___hrs

RECEIVING TECH/STAFF ...................................................................... Date /Time received: Date_____/______/__________Time ____: _____hrs

Comment: ..........................................................................................................................................................................................
