## Supplementary material for "A user-centred approach to developing a digital laboratory information system for high-volume clinical data: lessons learned from a malaria study in Kenya": SI Annex 2

### Subject Microscopy Summary

*View subject microscopy Summary*

Bloodslide Code / Subject ID

Study Phase: Cohort 1

Reader Code: All

Status: All

Bloodslide Date Taken: No date selected

Review Status: All

Urgent Status: All

Total Results: 27958

| Bloodslide Code | Subject ID | Date | Read 1 | Read 2 | Read 3 | Final MCR Result | Final Species ID | Final Parasite Density |
|---|---|---|---|---|---|---|---|---|
| [illegible] | [illegible] | 2021-10-26 | negative | negative | | negative | None | |
| [illegible] | [illegible] | 2021-10-26 | positive | positive | | positive | Pf | 671.9166614990285 |
| [illegible] | [illegible] | 2021-10-26 | negative | negative | | negative | None | |
| [illegible] | [illegible] | 2021-10-26 | negative | negative | | negative | None | |
| [illegible] | [illegible] | 2021-10-26 | negative | negative | | negative | None | |
| [illegible] | [illegible] | 2021-10-26 | positive | negative | positive | positive | Pf | 369.08535598151275 |
| [illegible] | [illegible] | 2021-10-26 | negative | negative | | negative | None | |
| [illegible] | [illegible] | 2021-10-26 | negative | negative | | negative | None | |

© 2023 — **#techsense**
